# Prevalence and Factors Associated with Cryptosporidiosis Among Livestock and Dogs in Kasese District, Uganda: A Cross-Sectional Study

**DOI:** 10.1101/2023.02.27.23286549

**Authors:** Clovice Kankya, Justine Okello, James Natweta Baguma, Rogers Wambi, Lesley Rose Ninsiima, Methodius Tubihemukama, Christine Tricia Kulabako, Musso Munyeme, Sonja Hartnack, Walter Okello, James Bugeza, James Muleme

**Author notes:** Corresponding author; Clovice Kankya.

## Abstract

**Background:** Cryptosporidiosis is a common cause of diarrheal disease in livestock and dogs, and it can result in significant economic losses due to decreased productivity and higher treatment costs. The cryptosporidiosis burden in livestock and dogs is largely unknown and underexploited. We designed a cross-sectional research study to determine the prevalence and factors associated with cryptosporidiosis in livestock and dogs.

**Methods:** The questionnaire data was downloaded from the Kobotoolbox server in excel format for cleaning and analysis. Laboratory results were matched with the questionnaire data and statistical analysis was performed using STATA version 14 and R version 4.2.2. Descriptive statistics were conducted to determine the frequencies, percentages, and proportions of the different study variables. For the categorical variables, 95% binomial and multinomial confidence intervals were obtained using the commands BinomCI(), with Jeffreys approach, and MultinomCI() available in the DescTools package. To adjust for potential clustering within household, a logistic mixed model approach with household as random effect was chosen and performed using the package glmmTMB. The outcome was a positive or negative test result of each dog. A likelihood ratio test using the lmtest package was used to determine if a predictor was significantly associated with the outcome. To adjust for multiple comparisons, the multcomp package with Tukeys approach was used. First all variables were included in a univariable approach. Second all variables with p-values <0.2 were included in a multivariable model..

**Results:** Dogs were over 50times more likely to have the cryptosporidium infection compared to goat. With aOR 56.07, 95%CI= [10.2569,306.5307] and p-value <0.001. Furthermore, being of Mukonjo tribe was over 20times more likely to have cryptosporidiosis compared to being Musongora. At aOR 24.92, 95%CI= [3.6971,168.0655], p-value 0.0010. Additionally, persons who drunk water from the river ponds were 76.93 times more likely to have cryptosporidium infection compared to the counterparts whose source of drinking water were from protected sources with aOR 76.93, 95% CI= [3.71,1595.324] p-value= 0.0050.

**Conclusion:** Cryptosporidiosis still remains a big public health problem in Kasese District. Dogs are more at risk of having cryptosporidiosis whereas drinking from river ponds was highly attributed to acquiring cryptosporidiosis.

## Introduction

Cryptosporidiosis is a diarrheal disease caused by the parasite Cryptosporidium. It is found worldwide and can affect a variety of animals, including humans, livestock, and pets (Pumipuntu & Piratae, 2018 and Ramirez et al., 2004). Cryptosporidium is more prevalent in developing countries (5% or higher) than in developed countries (3% or less) (Starkey et al., 2006). In humans, the disease is typically transmitted through the ingestion of contaminated food or water or through close contact with infected individuals or animals (Barnes et al., 2017 and Ganter, 2015). It can cause symptoms such as diarrhea, abdominal cramps, and dehydration, which can be particularly severe in people with weakened immune systems, such as those with HIV/AIDS or cancer (Xiao & Griffiths, 2020). Recently, there has been a documented increase in opportunistic infections and risk of acquiring opportunistic cryptosporidium infections in humans with HIV/AIDS patients and other co-morbidities in Kasese district (Kankya et al., 2022).

In livestock, cryptosporidiosis can lead to weight loss, reduced milk production, and decreased growth rates, all of which can have significant economic impacts for farmers. The disease can also be transmitted to humans through the consumption of contaminated meat or milk. Cryptosporidiosis can affect a variety of animal species, including cattle, sheep, goats, pigs, dogs and poultry (Smith et al., 2021). Dogs can be infected with Cryptosporidium and may shed the parasite in their faeces, which can contaminate the environment and potentially transmit the infection to humans (Ahmed et al., 2021).

In Uganda, cryptosporidiosis in calves and lambs are considered as the main source for human cryptosporidiosis, like in other low-income countries. With the increasing number of farming systems in Kasese district and Uganda at large, cryptosporidiosis in livestock is becoming a significant problem for animal health (both subclinical and clinical) and economic losses because of increasing need for veterinary services and labor costs due to increasing animal health-care cost with ultimate decreased growth rate of animals and mortality associated with severely affected animals. It is important to note that Cryptosporidium oocysts excreted faeces from infected farm animals, particularly calves, can be a source of human infection and may have a great influence on public health (Starkey et al., 2006). The habitual occurrence of floods in Kasese district also increases the risk of transmission of cryptosporidium among humans and animals. Flood water can contain sewage and other contaminants that may be contaminated with Cryptosporidium oocysts that when humans and animals encounter, they may become infected with the parasite. In addition, flooding can disrupt water treatment and distribution systems, leading to the contamination of drinking water with Cryptosporidium oocysts. Flooding can also lead to the displacement of people and their animals, which can increase the risk of disease transmission through overcrowding and a lack of proper hygiene facilities. All these scenarios have been document in Kasese district (Boyce et al., 2016), hence a strong justification for selecting the study area.

Despite several infection control efforts such as increasing access to clean water and improving the quality of water sources, improving hygiene and sanitation, treating, and preventing malnutrition, Cryptosporidium infection remains a huge challenge in Kasese district of Uganda. Unfortunately, there has been no published information on the epidemiology of cryptosporidium infection in this area. This study therefore sought to fill the knowledge gap by determining the prevalence and factors associated with cryptosporidium in livestock and dogs in Kasese district.

## Materials and methods

### Study design

The study employed a cross-sectional study design with multistage sampling. It was conducted in Kasese district in Western Uganda from September 2019 to February 2020. The study was conducted among households with livestock and dogs.

### Sample size determination

Sample size determination was based on the previous studies on prevalence and associated risk factors in the East African region (Tombang et al., 2019). Based on the simple assumption that an absolute minimum of 50 domestic animals with cryptosporidium infection in the study population, we opted for 733 as a necessary minimum sample size, enabling also a reliable comparison between population factors at a prevalence of 20% Ausvet Epitools (Sergeant, 2018). With the existing logistics, facilities and specified period, a total of 89 household heads were interviewed and a total of 785 animal faecal samples were collected from three Karusandara, Katwe-Kabatooro town council and Lake Katwe Sub-county respectively.

### Study area and study population

The study was carried out in Kasese district of Uganda. Kasese is in the western region of Uganda, bordering Bunyangabu District in the East, Kitagwenda in the Southeast by Lake George, Rubirizi District in the South, the West by the Democratic Republic of Congo and Bundibugyo District in the North. Administratively, Kasese district is comprised of 27 Sub-Counties, 14 Town Councils, 1 Municipality with 3 Divisions. It has two vast counties of Bukonzo and Busongora with 5 Constituencies. It lies between latitudes 0° 12’S and 0° 26’N; longitudes 29° 42’E and 30° 18’E (Karyeija & Kahika, 2017). Kasese district has a population of 694,987 people and a total of 139,406 households (UBOS, 2016). Kasese is a multi-ethnic district with many people of different ethnic backgrounds. The main languages and ethnic groups that dominate the area are the Bakonzo, Basongora, Banyabindi, Batooro, Banyankole, and Bakiga respectively. The district is predominantly agricultural with livelihoods relying mainly on farming. The inhabitants also carry out fishing and trading.

The district has a considerably high animal population, estimated at 101,453 cattle, 300,518 goats (Benson & Mugarura, 2013). Majority of the farmers keep cattle, goats and dogs with close interaction which is a significant risk factor for transmission of various disease agents including cryptosporidium among the susceptible hosts.

### Sampling strategy and study participants recruitment

Our study used a multi staged sampling approach where sub counties, parishes, and villages were randomly selected using a random number generator. Karusandara, Katwe-Kabatooro town council and Lake Katwe Sub-county were selected for this study. For the household selection, we used a passive sampling method based on the presence of livestock. The recruitment team included a local veterinarian, research assistant, and a member of the local council from the study site. From each selected household, we sampled all the cattle, goats, and dogs present.

### Faecal sample collection

Faecal samples were collected from the rectum of goats, cattle, and dogs using sterile arm-length gloves. Faecal sample collection was conducted by a trained veterinarian attached to the study site. The samples were placed in sterile ziplock bags and stored at -4 degree celsius until they could be transported in cool box containing ice packs to the microbiology laboratory at the College of Veterinary Medicine, Animal Resources and Biosecurity. The samples were handled with care to ensure their integrity and maintain the accuracy of the results.

### Questionnaire data collection

To gather information on household, animals, and key activity characteristics, we administered pre-tested semi-structured questionnaires to the heads of households. The questionnaire designed in an online platform (Kobotoolbox) was used to collect data on a variety of factors, including the social demographic characteristics of the household heads, livestock husbandry practices, the sources of water used by both humans and animals, and the livestock treatment practices employed by the households. To ensure the accuracy and reliability of the collected data, these questionnaires were developed based on a review of relevant literature and with input from experts in the field and were pre-tested in Muhokya Sub-county in the same district before being used for final data collection. In addition to collecting data from the households, we also obtained data on the individual animals from which faecal samples were collected using an animal characteristics checklist. These two sets of data were later merged during the data curation and analysis process to provide a comprehensive overview of the study population and the factors associated with the presence of cryptosporidium.

### Laboratory procedures Sample reception

Upon arriving at the laboratory, the samples were assigned laboratory identification numbers and entered to the sample registry. To ensure accurate analysis, only samples that met certain criteria were accepted. These criteria included proper labeling, leak-proof containers, appropriate sample volumes, clean sample collection containers, and relevant sample information. Any samples that did not meet these requirements were immediately rejected and recorded in sample rejection forms

### Sample analysis

Faecal samples were processed for Cryptosporidium detection using the modified Ziehl-Neelsen (MZN) staining technique (Tombang et al., 2019). Briefly, a direct faecal smear was prepared on a clean, grease-free glass slide and allowed to air dry. The smear was then flooded with methanol for five minutes, air-dried, and briefly flame-fixed. The slide was then flooded with concentrated Carbol fuchsin and allowed to stain for 40 minutes. After washing with running tap water for 5 minutes, the slide was decolorized using 10% Suphuric acid for 15 seconds and washed again with running tap water. The slide was counter stained with 5% Malachite green for 5 minutes, washed with running tap water for 5 minutes, and allowed to air dry. The slide was then examined under a light microscope at x40 and x100 magnification for the presence of Cryptosporidium oocysts.

### Data analysis and management

The questionnaire data was downloaded from the Kobotoolbox server in excel format for cleaning and analysis. Laboratory results were matched with the questionnaire data and statistical analysis was performed using STATA version 14 and R version 4.2.2. Descriptive statistics were conducted to determine the frequencies, percentages, and proportions of the different study variables. For the categorical variables, 95% binomial and multinomial confidence intervals were obtained using the commands BinomCI(), with Jeffreys approach, and MultinomCI() available in the DescTools package. To adjust for potential clustering within household, a logistic mixed model approach with household as random effect was chosen and performed using the package glmmTMB. The outcome was a positive or negative test result of each dog. The predictor variables included:

A likelihood ratio test using the lmtest package was used to determine if a predictor was significantly associated with the outcome. To adjust for multiple comparisons, the multcomp package with Tukeys approach was used.

First all variables were included in a univariable approach. Second all variables with p-values <0.2 were included in a multivariable model.

### Quality control and assurance

All samples were verified to confirm the labeling, type, adequate volume, and integrity at the laboratory. A trained veterinarian collected animal faecal samples during sample and data collection. The sample containers were labelled with the unique codes that represented the household and animal type. All the samples were handled as highly infectious and personal protective equipment were used all times whenever any activity was to take place on the samples.

## Results

The study recruited 89 households in Kasese district, from which livestock were selected and questionnaires were administered. Specifically, 32, 16 and 41 households were in Karusandara, Katwe-Kabatooro town council and Lake Katwe Sub-County respectively.

**Table 1. showing Household characteristics of study respondents**

In the present study, 34.38% of the female animals were positive for the cryptosporidium infection unlike the male counterpart in whom the prevalence of cryptosporidiosis is 44.26%. Among the animals that showed signs of the disease, 32.9% had cryptosporidium infection. The prevalence of cryptosporidium infection was 41.82% among animals that were not treated for helminth unlike their counterparts that were treated for treated that had 14.76%.

### Prevalence of cryptosporidium

Overall prevalence of Cryptosporidium in Livestock and dogs in Kasese district was 36.7% (288/784). Specifically, the prevalence was 72.0% (18/25), 39.3% (217/552) and 25.5% (53/208) in Dogs, Cattle, and Goats respectively as shown in figure 2 below.

**Figure 1.**
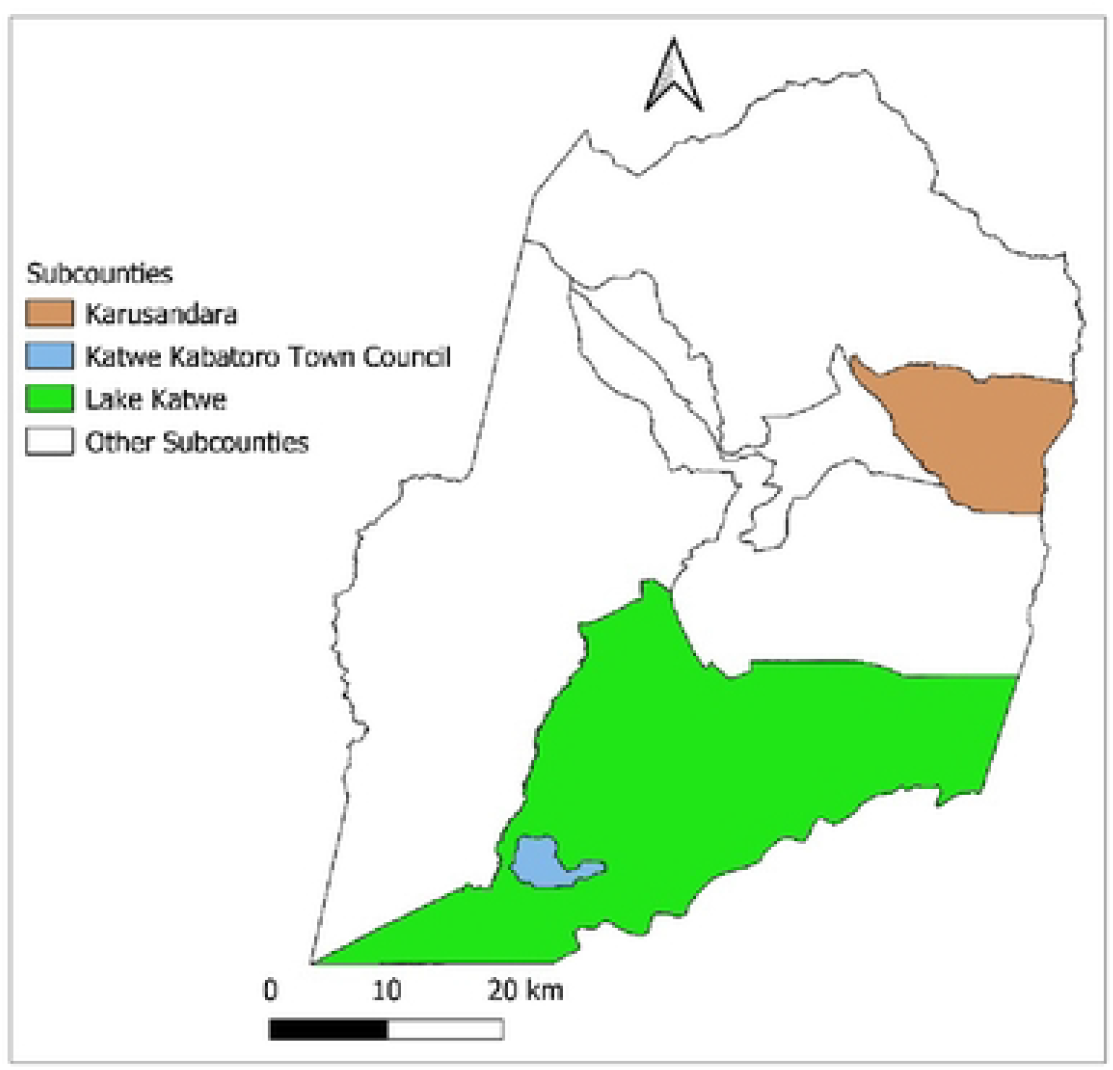
showing the study area.

**Figure2:**
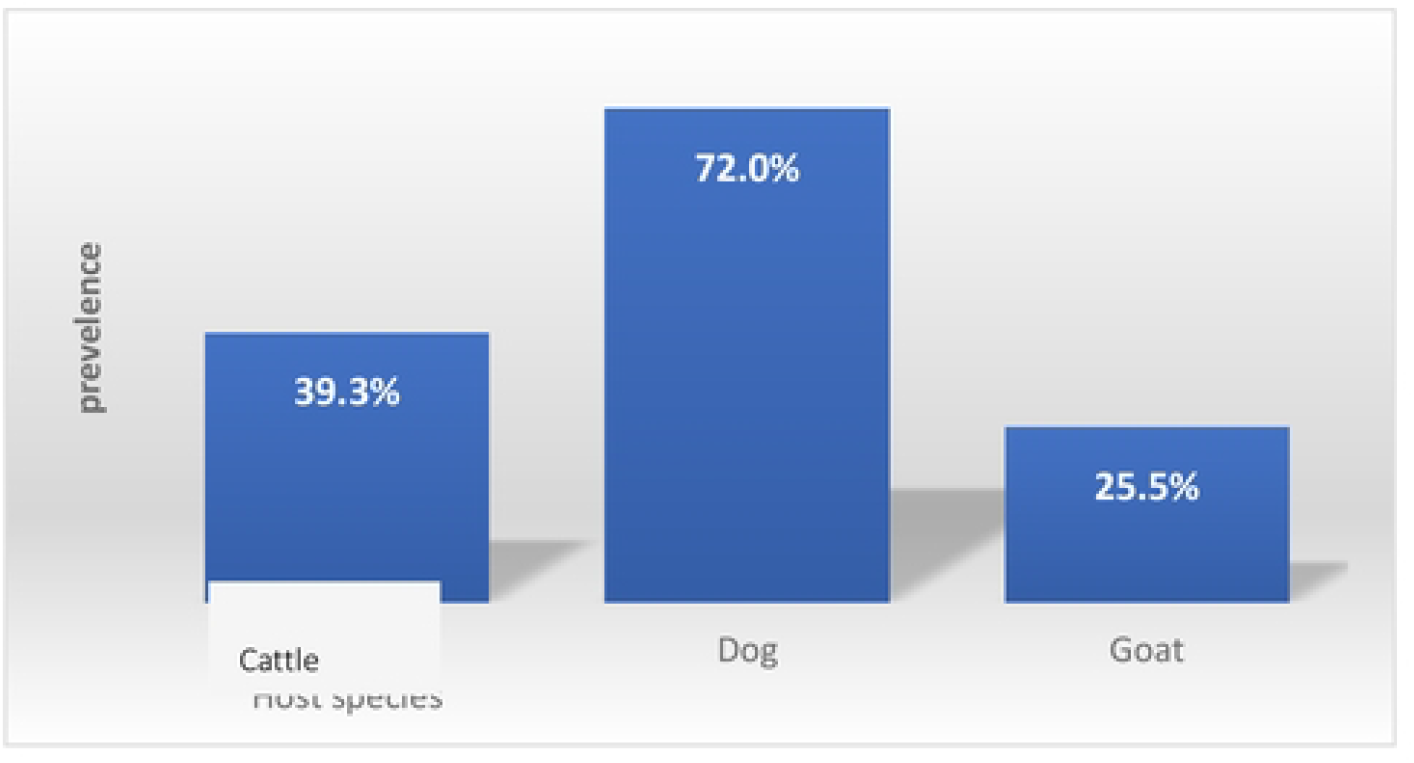
Showing prevalence of cryptosporidiosis in dogs, cattle, and goats.

### Risk factors associated with the prevalence of cryptosporidiosis

For the herd factors, Male animals were 1.47 times more likely to have cryptosporidiosis compared to the female counter parts. At aOR 1.4747, 95%CI= [0.8214,2.6473] and p-value 0.19, this association was however not statistically significant. Also, dogs were over 50times more likely to have the cryptosporidium infection compared to goat. With aOR 56.07, 95%CI= [10.2569,306.5307] and p-value<0.001; this association was statistically significant. Young animals were 1.03 times more likely to have cryptosporidiosis compared to their adult counterparts. At aOR 1.038, 95%CI= [0.5776,1.8654] and p-value 0.90; this association was however not statistically significant.

Furthermore, for human factors; being of Mukonjo tribe was over 20times more likely to have cryptosporidiosis compared to being Musongora. At aOR 24.92, 95%CI= [3.6971,168.0655], this association was statistically significant at p-value 0.0010. in addition, for water factors persons who drunk water from the river ponds were 76.93 times more likely to have cryptosporidium infection compared to the counterparts whose source of drinking water were from protected sources. At aOR 76.93, 95% CI= [3.71,1595.324], this association was statistically significant at p-value 0.0050.

**Table 2: Showing the risk factors associated with the cryptosporidium infection**

## Discussion

The present study focused on the diagnosis of Cryptosporidium spp. in livestock and dogs in the farming rural community set-up using modified Ziehl–Neelsen technique. The overall prevalence of Cryptosporidium in this study was 36.7% and this was much lower than the 70 % prevalence reported by (Romero-Salas et al., 2016). On the other hand, the current study reported a high prevalence than the 16% prevalence reported in Tanzania by Mtambo et al. The high prevalence reported in the present could have been attributed to the fact that most of the animals sampled in this study were kept under free range system. This system is predominant in agro-pastoral communities and has been linked to increased risk of exposure of animals to infectious agents including cryptosporidium (Nakayima et al., 2019). The current study revealed highest prevalence of cryptosporidium in dogs followed by goats and least in Cattle. This trend could be associated with different exposure levels among the animal species in the study. The highest prevalence in dogs in this study could be attribute to the fact that most of the dogs kept in this area are freely roaming and to some extent are likely to cross to the neighboring Queen Elizabeth national park which increases risk of exposure to infectious pathogens (Kankya *et al*., 2022).

Our present study reveals that male animals are more likely to have cryptosporidium compared to their female counterpart. Our study is in disagreement with a study by (Li *et al*., 2022) that reported a higher global prevalence and risk factors for cryptosporidium in female equus compared to the male. The high likelihood of the male animals having cryptosporidiosis in our study could be attributed to their behaviors like reproduction among others that provides fertile environment for the pathogens to attach onto the animal. For example, the male animals often mounts many female other animals and also grazes sometimes far from the grazing field which further exposures them to come into contact with infectious pathogens like cryptosporidium.

Furthermore, our study revealed that animals which drink water from river ponds were more likely to have cryptosporidium infection compared to those which drink water from protected springs and rainwater/valley dams. Our study is in perfect agreement with the study conducted by (Khan et al., 2019) who found that surface waters such as springs, river ponds among others are at higher risk of spreading cryptosporidium pathogen. It is imperative to note that such surface waters in the study area are likely to act as wallowing grounds for both domestic animals and wild animals which ultimately increase the risk of infection spread to those animals that come to drink such waters. Also, the high risk of acquiring cryptosporidium in this study could be attributed to the fact that protected springs and valley dams provides an optimum environment for the survival and multiplication of the cryptosporidium pathogen. This therefore is easily picked by the animals that encounters such waters during the drinking process. Also, the low risk of the protected waters in the spread of cryptosporidium infection to the animals in our study could be attributed the fact that such waters are always highly protected from almost all infectious pathogens that can infect animals and humans. And this stops other infectious pathogens from taking advantage of such water sources.

Our study further shows that animals that were regurlarly treated for the helminth infestations were at low risk of acquiring the cryptosporidium infection. Our finding is in agreement with a study by (Liao et al., 2020) who indicated that deworming was a protective factor against cryptosporidiosis. This could be attributed to the fact that different antihelminth drugs that are often administered to the animals exerts a static (limiting growth) and/or cidal (killing) effects on the cryptosporidium pathogen. The antihelminth treatment thus reduces the risk of the cryptosporidiosis in livestock and dogs.

Also, the animal in the present study, that presented with other underlying ailment were more likely to be infected with cryptosporidium. This finding were in agreement with a study carried out by (Ebiyo & Haile, 2022). This could be explained by the fact that underlying ailment has immunity lowering effect that paves way for establishment of cryptosporidium infection. Therefore, animals with underling ailment should be investigated and treated for cryptosporidium infection.

Our study acknowledges the limitations experienced during the execution of this study. Specifically, the distance between the diagnostic laboratory and study area may have affected the viability of cryptosporidium oocysts in samples with low faecal parasites hence undetectable and the observed prevalence. Also, we could not confirm the species and genotypes of Cryptosporidium based on the method employed in this study.

## Conclusion

The presents study indicates a high prevalence of cryptosporidiosis in Kasese district. The prevalence was further different cross the animal species examined with Dogs having the highest prevalence, followed by Cattle and goats. Being of male gender in the animals, drinking water from river ponds as well as the age (young) of the animals were associated with the risk of cryptosporidiosis infection. Regularly treating the animals for helminth infestation was a protective factor against cryptosporidiosis infection.

### Recommendations

- More efforts such as fencing of the surface water sources like spring waters should be effectuated to reduce the risk of exposure of the cryptosporidium infection at the Animal-Human-Environment interface.
- Also, community animal owners should take initiatives to frequently treat intestinal infestations on their livestock and dogs such as deworming. This can be done by using both conventional drugs as well as locally available (herbal concoctions) therapy
- Furthermore, community members should always practice optimum hygiene such as hand washing, proper disposal of faecal matter as sustainable approach to prevention and control of cryptosporidiosis spread at the animal-human-environment interface.
- Additionally, all the concerned stakeholders including government, private sectors, farmers, academia among others will need to work together to improve the quality of animal husbandry practices within the agro-pastoral communities

## Data Availability

All data produced in the present study are available upon reasonable request to the authors
All data produced in the present work are contained in the manuscript

## Data availability statement

The data used in the generations of this manuscript will be made available to any individual upon request.

## Ethical approval

The authors sought ethical clearance and the research protocol was approved by the research ethics committee at the Makerere University School of Biosecurity, Biotechnical and Laboratory Sciences (SBLS) and the Uganda National Council for Science and Technology (UNCST) - (NS 640). The study participants provided their written informed consent to participate in this study.

We also followed the declaration of the Helsinki guidance on ethical principles involving humans subjects in this study.

## Acknowledgments

The authors extend sincere appreciations to the reviewers and editors of this manuscript to ensure its publication. Also, special thanks goes to the research assistants who worked tirelessly hard to support the data, and sample collection during the execution of this research study. We also thank the government of NORWAY, through the NORHED Ι programme that provided financial support used in this study.

